# SARS-CoV-2 oral tablet vaccination induces neutralizing mucosal IgA in a phase 1 open label trial

**DOI:** 10.1101/2022.07.16.22277601

**Authors:** Susan Johnson, Clarissa I Martinez, Clara B Jegede, Samanta Gutierrez, Mario Cortese, C Josefina Martinez, Shaily J Garg, Nadine Peinovich, Emery G Dora, Sean N Tucker

## Abstract

**Background:** Despite the plethora of efficacious vaccines to the initial Wuhan strain of SARS-CoV-2, these do not induce robust mucosal immunity, offering limited protection against breakthrough infection and replication in the respiratory tract. The mucosa is the first line of defense, therefore a vaccine that induces a mucosal IgA response could be an important strategy in curbing the global pandemic.

**Methods:** We conducted a single-site, dose-ranging, open-label clinical trial of an oral SARS-CoV-2 vaccine to determine safety and immunogenicity. This tablet vaccine is comprised of a non-replicating adenoviral vector expressing the SARS-CoV-2 Spike and Nucleocapsid genes and a double-stranded RNA adjuvant. 35 adult subjects meeting inclusion/exclusion criteria received a single low (1×10^10^ IU) or high (5×10^10^ IU) dose and 5 subjects received two low doses. Nasal, saliva and serum samples were assessed for the presence of IgA, IgG and surrogate neutralizing antibodies. Convalescent subjects between 1-8 months post infection were recruited to give nasal, saliva, and serum samples for comparison.

**Results:** The vaccine was well tolerated without any dose-limiting toxicity observed. No serum neutralizing antibodies were observed, but modest IgA responses were seen in serum post immunization. The majority of vaccine recipients had an increase in mucosal secretory IgA which was highly cross-reactive against all coronaviruses tested and persisted up to 360 days. Furthermore, the nasal IgA induced by vaccination has superior neutralizing activity compared to convalescent nasal samples.

**Conclusion:** The vaccine was safe, well tolerated and generated mucosal immune responses including cross-reactive surrogate neutralizing secretory IgA. These results demonstrate the ability of a mucosal vaccine to induce long-lasting mucosal IgA to SARS-CoV-2.

**Graphical Abstract:** 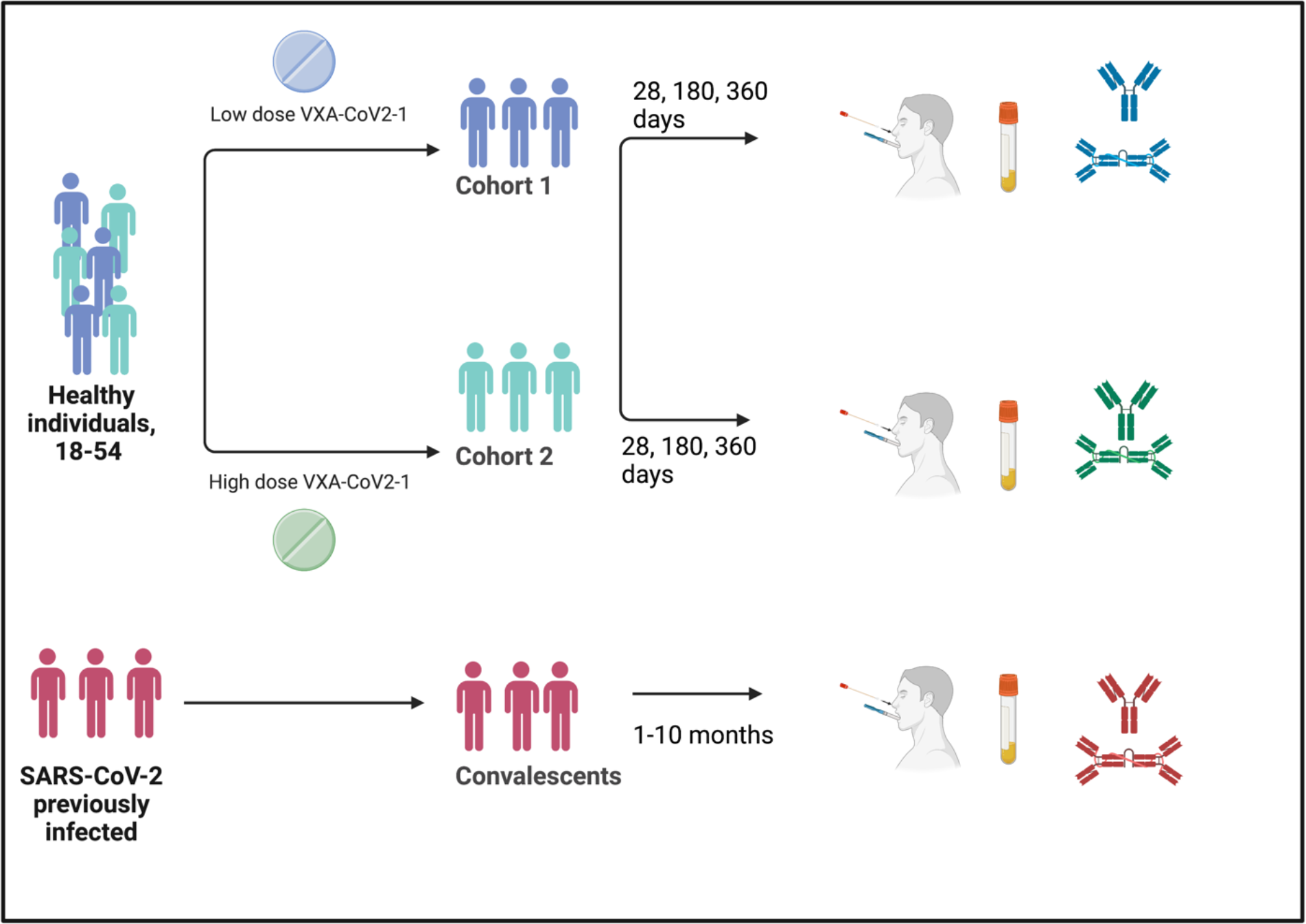

## Introduction

Vaccination has provided substantial health benefits to recipients during the global SARS-CoV-2 pandemic, and was critical to increasing survival and providing initial protection against symptomatic disease. However, as the SARS-CoV-2 virus has evolved, the current vaccines have been unable to curb large outbreaks of current SARS-CoV-2 variant strains such as omicron. As well as the health outcomes from repeated infections, this has had devastating effects on the economy, with the number of people out of work due to covid related reasons increased 32% during the peak of the first omicron variant (1). One of the main reasons for this is that the current SARS-CoV-2 parenteral vaccines might not provide the breadth of immunity needed at the site of infection causing breakthrough infections in the recipients, and allowing increased viral replication in the upper respiratory tract, leading to viral shedding, and transmission to other people. Further, viral evolution may lead to more severe consequences of infection, so there’s consensus that new vaccine strategies may be needed.

Vaxart has developed a shelf-stable oral tablet vaccine that incorporates both the spike (S) and nucleocapsid (N) proteins. Vaxart’s vaccine platform uses a non-replicating Adenovirus type 5 (Ad5) backbone and a TLR3 agonist as an adjuvant. These vaccines spanning different indications have been administered to over 650 subjects, have been well tolerated, and previously able to generate robust humoral and cellular immune responses to the expressed antigens (2, 3).

Our oral vaccine platform increases adaptive immune responses at mucosal surfaces, which are important sites of entry for respiratory pathogens such as SARS-CoV-2 and potentially important sites for long-term low level viral replication of SARS-CoV-2 (4). Mucosal IgA, due to its polymeric nature and ability to translocate, is also more likely to hinder transmission (5) (6) (7); a vaccine that could decrease asymptomatic transmission could alleviate outbreaks. Dimeric IgA, such as found in the mucosa, has been shown to have enhanced neutralization capacity against SARS-CoV-2 *in vitro* (8) and can be cross-reactive against viral variants (9). Secretory IgA has been detected in the milk of SARS-CoV-2 infected mothers and can be passed onto the neonate, indicating that secretory IgA mediated protection could be transferred to neonates prior to being eligible for vaccination (10).

Here we describe the results of our clinical trial testing Vaxart’s oral SARS-CoV-2 vaccine candidate. The objectives of the single center, open-label, dose-ranging Phase 1 clinical trial were to determine the safety and immunogenicity of two dose levels of the oral COVID-19 vaccine candidate, termed VXA-CoV2-1, in normal healthy subjects.

## Results

### Demographics

Between September and November 2020 at a single site in Cypress, CA, a total of 66 subjects were screened, and 35 healthy adult volunteers aged 18-53 years old who tested negative for SARS-CoV-2, via rapid antigen test and confirmatory PCR, were enrolled to receive either a single dose or a two-dose vaccination regimen of VXA-CoV2-1. Subjects were enrolled into the study in three cohorts; a sentinel low dose with a boost at Day 29 (Cohort 1), a single low dose (Cohort 2) and a single high dose (Cohort 3) (Figure 1). Subject demographics are described in table 1. The average age of study participants was 34.2 years with good balance between the mean age of low (35.2 yrs) and high dose (32.7 yrs) cohorts. The study enrolled a higher percentage of male participants (23; 65.7%) compared to female participants (12; 34.3%).

**Figure 1.**
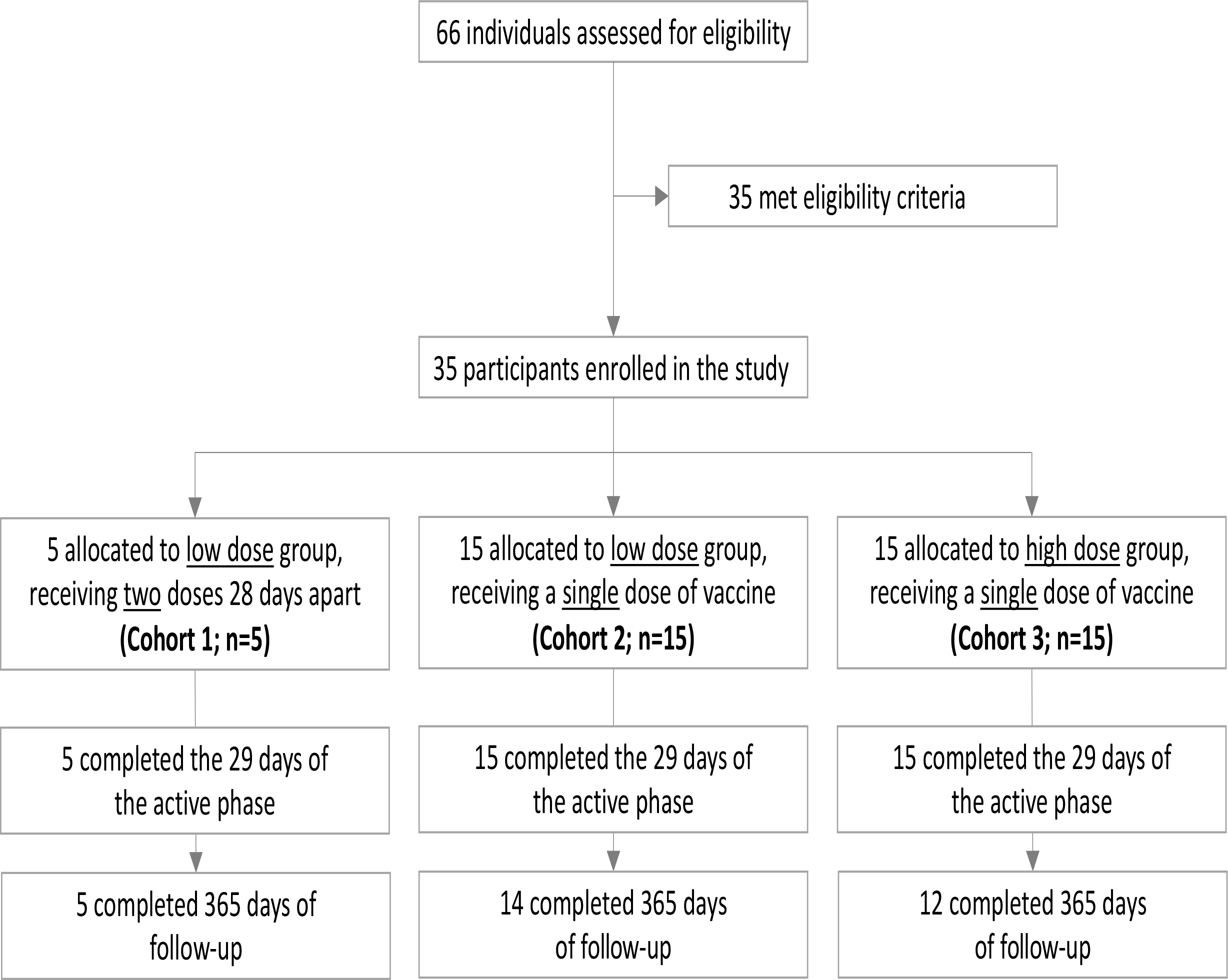
Oral vaccine VXA-CoV2-1 clinical trial profile.

**Table 1.**

| Characteristics | Cohort 1 (N=5) | Cohort 2 (N=15) | Cohort 3 (N=15) |
| --- | --- | --- | --- |
| Age (years) |  |  |  |
| Mean (SD) | 40.4 (10.24) | 33.5 (11.05) | 32.7 (8.98) |
| Median | 43.0 | 32.0 | 31.0 |
| Min - Max | 25 - 51 | 18 - 53 | 21 - 52 |
| Sex |  |  |  |
| Male | 4 (80.0%) | 8 (53.3%) | 11 (73.3%) |
| Female | 1 (20.0%) | 7 (46.7%) | 4 (26.7%) |
| Race |  |  |  |
| White | 3 (60.0%) | 9 (60.0%) | 8 (53.3%) |
| Black or African American | 1 (20.0%) | 3 (20.0%) | 3 (20.0%) |
| Asian |  |  |  |
| American Indian or Alaska Native |  |  |  |
| Native Hawaiian or Other Pacific Islander | 1 (20.0%) | 3 (20.0%) | 1 (6.7%) |
| Other | 0 (0.0%) | 0 (0.0%) | 3 (20.0%) |
| Ethnicity |  |  |  |
| Hispanic or Latino | 2 (40.0%) | 3 (20.0%) | 5 (33.3%) |
| Not Hispanic or Latino | 3 (60.0%) | 12 (80.0%) | 10 (66.7%) |
| BMI (kg/m²) |  |  |  |
| Mean (SD) | 26.3 (2.2) | 25.9 (2.7) | 24.4 (3.3) |
| Median | 26.4 | 27.2 | 24.3 |
| Min - Max | 23.1 - 28.4 | 19.4 - 29.4 | 19.9 - 28.8 |

SD = standard deviation; kg = kilogram; m² = square meter

### Summary of adverse events

The primary endpoint of the study was safety and tolerability with a focus on frequency of AEs of reactogenicity for one week post vaccination (solicited symptoms through Day 8) as well as any unsolicited AEs reported through 28 days post last vaccination, Day 57 for Cohort 1 and Day 29 for Cohorts 2 and 3. Long term safety through 12 months post vaccination included monitoring for serious AEs (SAEs), medically attended AEs (MAAEs), and COVID-19. A summary of subjects with solicited AEs collected for 7 days after each dosing is presented in Table 2. A total of 13 subjects (37%) from Cohorts 1, 2, and 3 experienced solicited AEs. Solicited symptoms of reactogenicity included fever, headache, myalgia, abdominal pain, anorexia, nausea, vomiting, diarrhea, and malaise. In the low dose groups (Cohorts 1 and 2) 4 of 20 subjects (20%) reported solicited AEs; in the high dose group (Cohort 3) 9 of 15 subjects (60%) reported solicited AEs. No subjects in Cohort 1 reported any solicited AEs after they received a second vaccine dose on Day 29. All solicited AEs were of mild or moderate severity, and no severe or life-threatening solicited AEs were reported during the study. No subjects discontinued on study due to solicited AEs and no concomitant medications were needed to treat any solicited AEs. Five subjects (14%) experienced unsolicited AEs through Day 57. All unsolicited AEs were reported in Cohorts 2 and 3, and all AEs were graded as mild. Two subjects had unsolicited AEs considered to be related to study treatment by the investigator. There were no SAEs, MAAEs or confirmed cases of COVID-19 reported through Day 57. All 35 subjects rolled into the Safety-Follow-up period where they continued to be monitored for these events through 12 months post first vaccination. During this period, 2 subjects withdrew consent, and 2 subjects were lost to follow-up. No subjects reported any SAE or MAAE during the Safety-Follow-up period. Two subjects reported confirmed COVID-19 infections, neither of which was a MAAE.

**Table 2.**

|  | Cohorts 1 & 2<br>- Day 1<br>(N=20) | Cohort 1<br>- Day 29<br>(N=5) | Cohort 3<br>- Day 1<br>(N=15) | Total -<br>Day 1<br>(N=35) | Total -<br>Days 1 and 29<br>(N=40) |
| --- | --- | --- | --- | --- | --- |
| Any Solicited AEs, n (%) | 4 (20.00%) | 0 (0.0%) | 9 (60.00%) | 13 (37.14%) | 13 (32.50%) |
| Fever | 0 (0.0%) | 0 (0.0%) | 2 (13.33%) | 2 (5.71%) | 2 (5.00%) |
| Headache | 3 (15.00%) | 0 (0.0%) | 1 (6.67%) | 4 (11.43%) | 4 (10.00%) |
| Myalgia (muscle pain) | 2 (10.00%) | 0 (0.0%) | 1 (6.67%) | 3 (8.57%) | 3 (7.50%) |
| Abdominal Pain | 0 (0.0%) | 0 (0.0%) | 2 (13.33%) | 2 (5.71%) | 2 (5.00%) |
| Anorexia (defined by not eating) | 0 (0.0%) | 0 (0.0%) | 2 (13.33%) | 2 (5.71%) | 2 (5.00%) |
| Nausea | 0 (0.0%) | 0 (0.0%) | 4 (26.67%) | 4 (11.43%) | 4 (10.00%) |
| Vomiting | 0 (0.0%) | 0 (0.0%) | 4 (26.67%) | 4 (11.43%) | 4 (10.00%) |
| Diarrhea | 0 (0.0%) | 0 (0.0%) | 4 (26.67%) | 4 (11.43%) | 4 (10.00%) |
| Malaise/Fatigue | 2 (10.00%) | 0 (0.0%) | 2 (13.33%) | 4 (11.43%) | 4 (10.00%) |

AE = Adverse Event

### VXA-CoV2-1 oral vaccination induces secretory mucosal IgA that persists up to one year later

Given the unique characteristics of the VXA-CoV2-1 oral vaccine candidate, we expected to observe SARS-CoV-2-specific IgA antibody responses in mucosal compartments.

Nasal samples were taken from subjects at Day 1 (baseline), 29, 180 and 360. At Day 29 post vaccination, 16/35 (46%) had a 1.5-fold or higher increase in secretory IgA over pre-vaccinated baseline, with half of those (8/16) showing a 2-fold higher increase (Figure 2A). Responses were observed to all 3 antigens tested, Spike, Nucleocapsid and RBD. 15/35 (43%) had a 1.5-fold or higher in SARS-CoV-2 specific nasal IgA that could bind to the delta or omicron spike variants (Figure 2B). While serum IgA increased in the vaccinated population, it was a mild increase with only 7/35 (20%) subjects were shown to have a 1.5 or above anti-SARS-CoV-2 IgA response in the serum (Figure 2C). Of the responders that had SARS-CoV-2 specific IgA, 5 of 11 (46%) still had nasal spike specific IgA responses 2-fold or above over pre-vaccination baseline with 3 subjects still having increased IgA one year later (Figure 2D). 6/11 subjects had RBD specific nasal IgA at day 180 with 3 subjects showing increased IgA at Day 360 (Figure 2E). 3/11 (28%) subjects with N specific IgA had persisting IgA at Days 180 and 360 (Figure 2F). Samples from the remaining responding subjects were not able to be obtained. Similar longevity of responses was seen in saliva, with 5/7 (72%) subjects having a 2-fold or above response (83%) against spike at 180 days post vaccination. Of these 5 subjects, 3 of them still had a 2-fold or above anti-spike response in the saliva one year post vaccination and 2 subjects had a 1.5-fold salivary anti-spike response (Figure 2G-I). 2/7 subjects had a salivary anti-RBD response at day 180, 2 subjects had two-fold or above anti-RBD response one year post vaccination and two had a 1.5-fold response at one year post vaccination. 8/15 subjects had a 2-fold or above response to N at Day 180 post vaccination, with six of these subjects still showing elevated levels of anti-N one year post vaccination.

**Figure 2:**
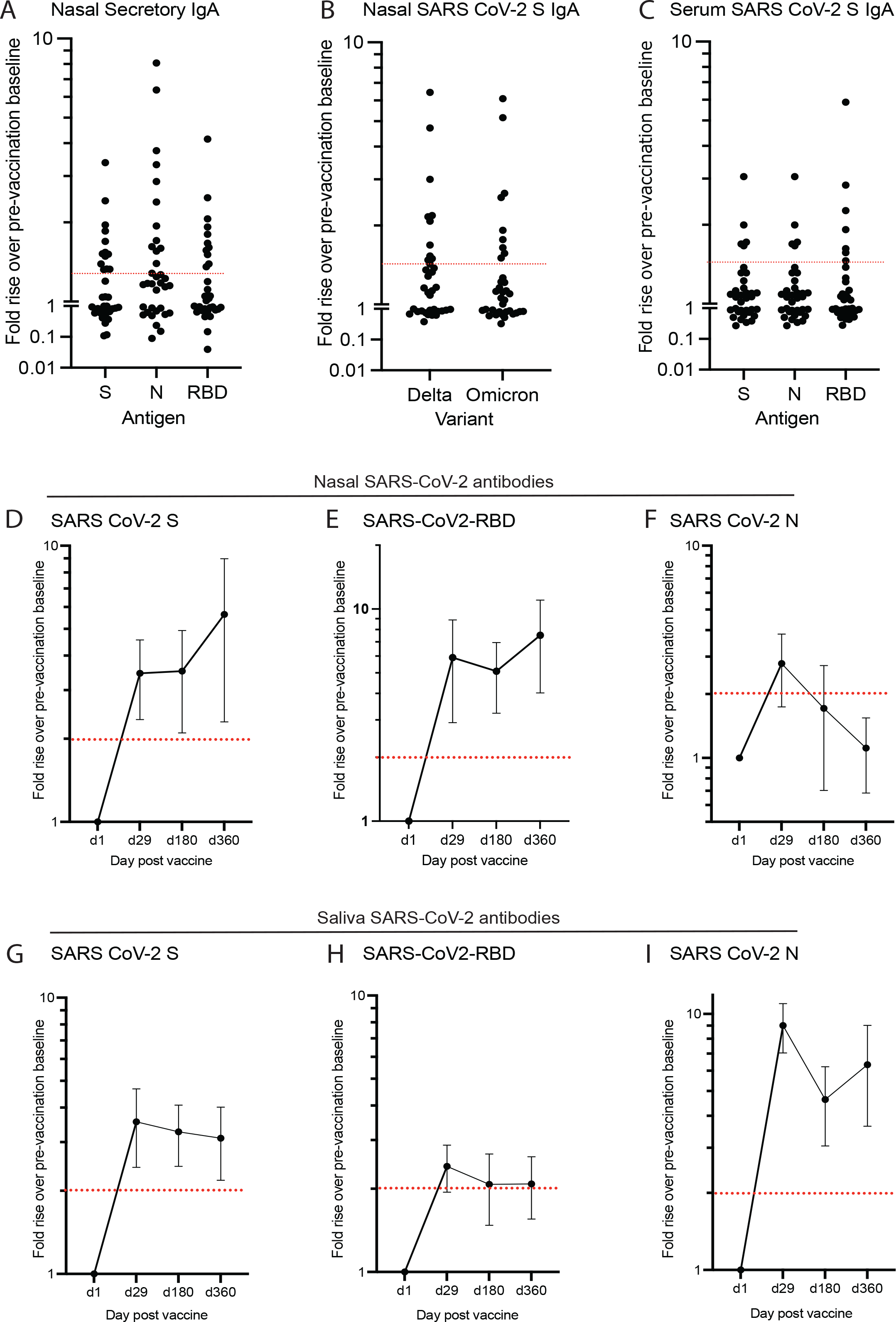
VXA-COV2-1 generates nasal and saliva IgA that is cross-reactive, neutralizing and can persist up to 180 days. IgA was measured using SARS-CoV-2 specific MSD assays. (A) Fold rise over baseline of secretory IgA in all subjects. (B) Fold rise over baseline of SARS-CoV-2 anti-spike IgA using delta or omicron variants in all subjects. (C) Fold rise over baseline of IgA in serum in all subjects. Red line represents 1.5 fold change. (D-F) Average of responders showing 50% of those who had an increase in S (D), RBD (E) or N (F) nasal IgA at d29 post vaccination maintained it 180 days post vaccination. Average of responders showing 50% of those who had an increase in S (G), RBD (H) or N (I) saliva IgA at d29 post vaccination maintained it 180 days post vaccination.

### VXA-CoV2-1 induces IgA in the mucosa to similar levels as convalescent subjects

Subjects not enrolled in the Vaxart tablet study, that had recovered from SARS-CoV-2 infection (between 1 and 10 months post infection) were recruited to donate sera, nasal swabs and saliva. IgA levels were measured via MSD and compared to the VXA-CoV2-1 subjects. As a whole, convalescent subjects showed a significant increase in SARS-CoV-2 spike specific nasal IgA but no difference in RBD or N-specific nasal IgA (Figure 3A-C). The levels of S and RBD-specific IgA was similar in saliva with a significant increase in N-specific IgA in VXA-CoV2-1 subjects (Figure 3A-C).

**Figure 3:**
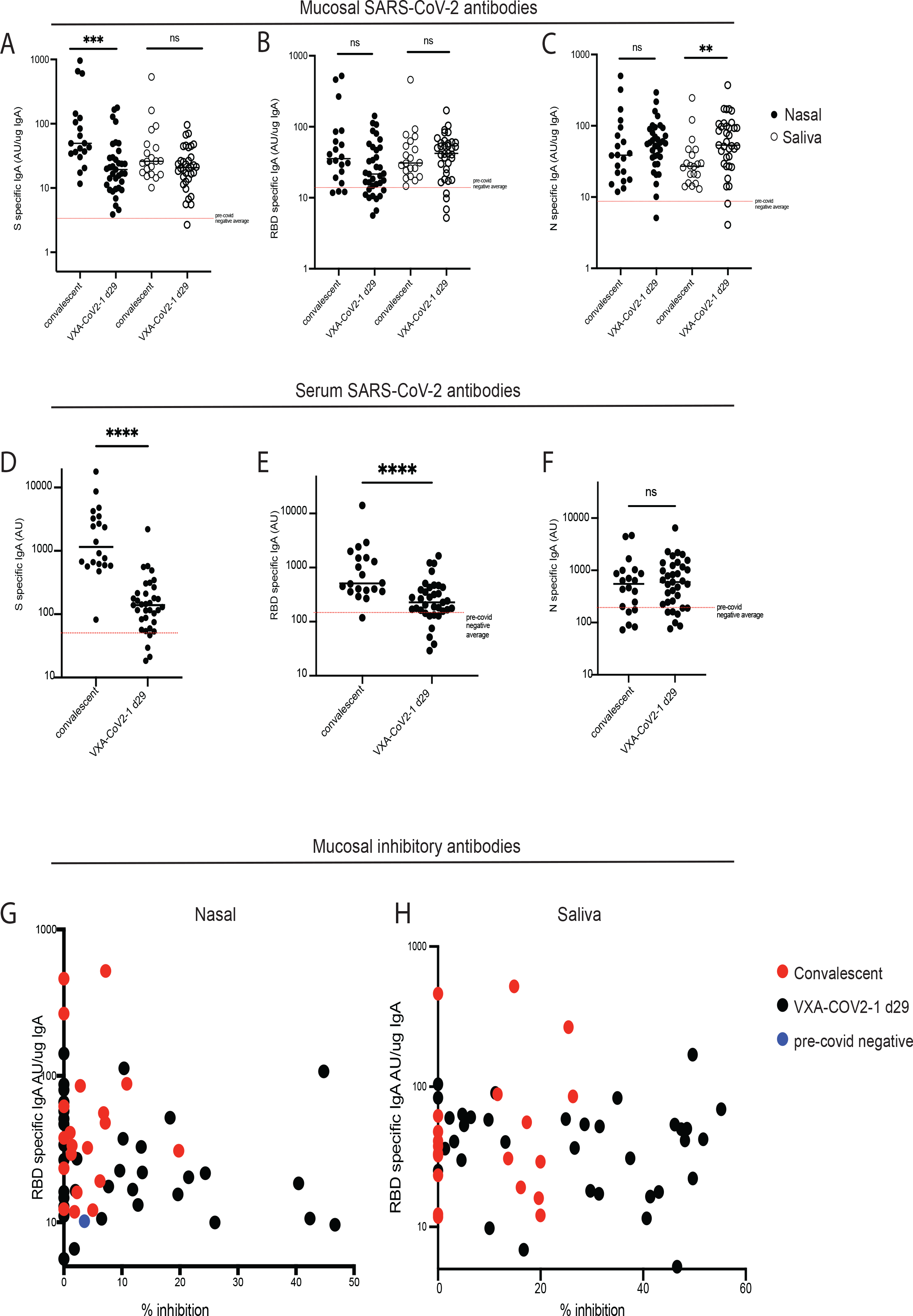
VXA-COV2-1 generates better neutralizing nasal and saliva antibodies than natural infection. (A-F) IgA in either convalescent or vaccinated subjects was measured using SARS-CoV-2 specific MSD assays. A-C shows mucosal antibodies and D-F shows serum antibodies. (G-H) Inhibition as measured via a surrogate neutralizing assay in nasal (G) or saliva (H), all subjects shown. Convalescent subjects in red, negative subject (pre-2019) shown in blue.

Significant differences between the convalescent group and the vaccinated group were seen in the spike and RBD specific IgA found in serum – with convalescents having a 10-fold increase over VXA-CoV2-1 trial subjects. No significant difference was seen in the N (Figure 3D-F).

### VXA-COV2-1 improves mucosal neutralizing IgA over convalescent individuals

Nasal and saliva samples were tested for their ability to inhibit binding of ACE-2 to RBD in a surrogate viral neutralizing test (sVNT). Inhibition and total amounts of RBD-binding IgA were correlated in several subjects, with the subjects that showed an above 2-fold increase in SARS-CoV-2 specific IgA having enhanced surrogate neutralizing activity (Figure 3G). When comparing the responses of natural infection vs oral vaccination, despite some convalescent subjects showing higher RBD specific IgA, 50% of the vaccinated cohort displayed more surrogate neutralizing activity than the convalescent subjects (Figure 3G). Similar responses were seen with the saliva samples, with the VXA-CoV2-1 subjects showing higher responses.

About 50% of this inhibitory activity in responders was maintained over time in both the nasal and saliva with most subjects having a higher surrogate neutralizing capacity 6 months post vaccination than pre-vaccination. Several subjects had an increase, most likely being boosted by natural exposure to SARS-CoV-2 or other coronaviruses (Figure 4A). Cross-inhibitory activity was also shown against SARS-CoV-1, with an average of 64% cross-inhibitory activity shown compared to the SARS-CoV-2 sVNT assay (Figure 4C).

**Figure 4:**
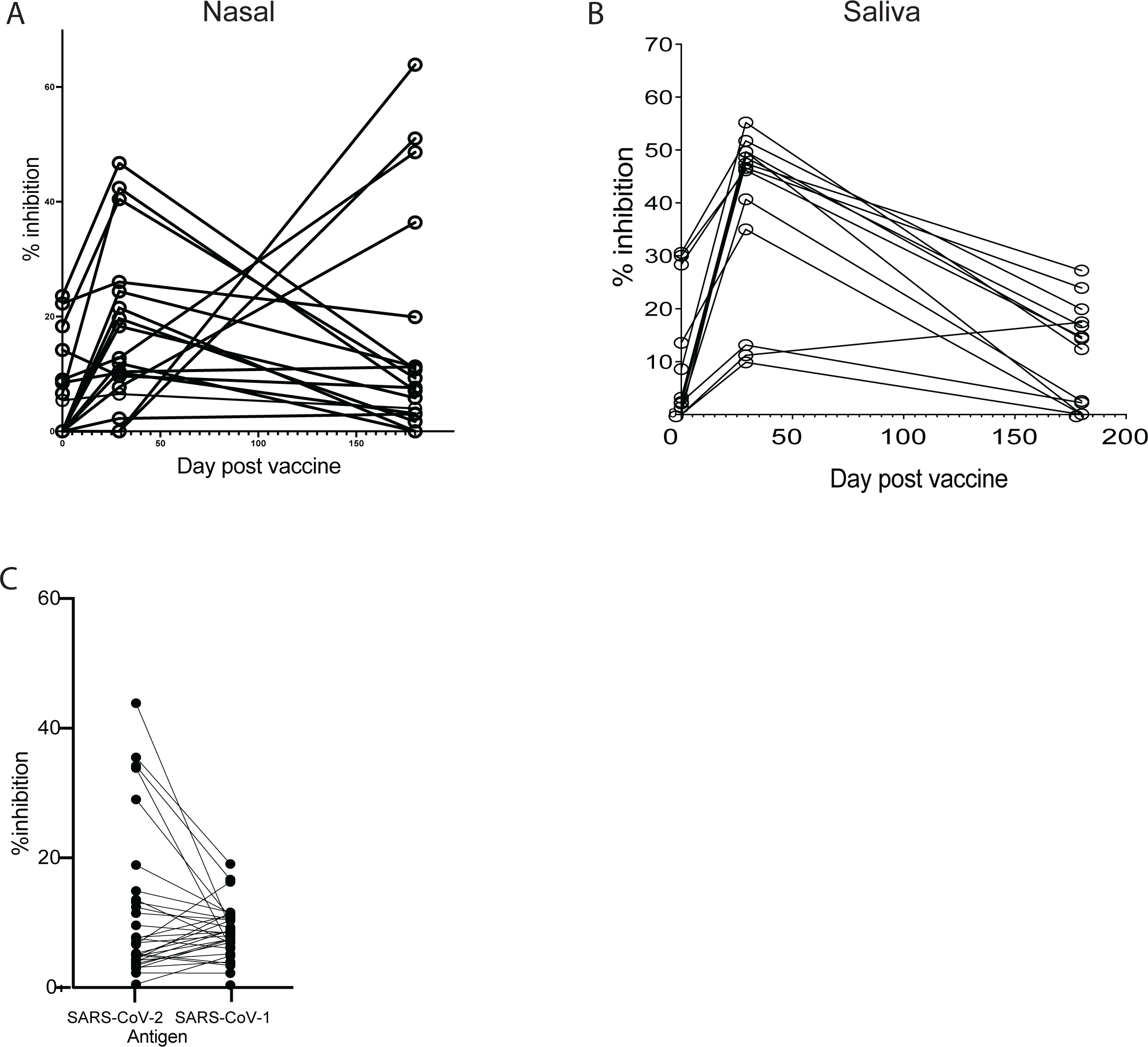
Neutralizing activity in mucosal samples is maintained over time and is cross-neutralizing. (A-B) Inhibition over time shown in nasal (A) or saliva (B) samples of responders, with the majority maintaining some degree of inhibition (C) Those that show SARS-CoV-2 inhibition in nasal samples at d180 also show SARS-CoV-1 inhibitory activity, with an average of 65% cross-reactivity.

### Additional Immune Assessments

No significant differences in vaccine-specific IgA responses in the saliva or nasal samples between the two dose groups could be observed (Supplementary Fig 1A, 1B). A significant increase in IgA+ β7+ B cells was seen at d29, indicating the presence of mucosal homing plasmablasts (supplementary figure 1C). No neutralizing antibody responses were found in the serum of any subject, and only 1 subject had a significant increase in IgG. Only small amounts of IgG above background were found in any of the nasal swabs, compared to the amounts of IgA (supplementary figure 2) with only three subjects having an above 2-fold increase. CD8 and CD4 T cell responses were induced in a 73% of subjects at day 8, with CD8 responses being particularly robust (manuscript in preparation).

## Discussion

In this study we demonstrated that our oral tablet vaccine candidate, VXA-COV2-1, was well tolerated and immunogenic in a clinical setting. Serum responses were low, but the mucosal IgA responses specific to SARS-CoV-2 S, N, and RBD increased in the majority of subjects. VXA-CoV2-1 generated cross-reactive secretory IgA that persisted past 180 days in the majority of responders after a single administration. SARS-CoV-2 specific IgA increased in both nasal and saliva secretions (11). Additionally, nasal IgA was shown to be secretory IgA, which has been shown to be superior to monomeric IgA at neutralizing SARS-CoV-2 *in vitro* (12). VXA-CoV2-1 induced similar levels of IgA in the nose and saliva as convalescent subjects. Furthermore, we observed that vaccine immunized subjects had improved mucosal neutralizing antibody responses compared to convalescent subjects, suggesting that vaccination might improve the quality of the antibody responses at mucosal surfaces. Further investigation into this phenomenon is required, but one possible explanation is that the SARS-CoV-2 virus misdirects a mucosal immune response, similar to what may occur with RSV infections (13).

The current most important issue facing vaccination campaigns, is that the parenteral vaccines elicit highly strain-specific responses to spike protein of the parental Wuhan strains, with lower cross-reactivity to the more recent delta and omicron variants. This has led to a large number of breakthrough infections. We found that vaccination with VXA-CoV2-1 elicited mucosal IgA that was highly cross-reactive to omicron and delta variants of SARS-CoV-2 as well as previously shown that both the saliva and nasal samples at day 29 post vaccination were cross-reactive to other human alpha and beta coronaviruses (11). In fact, the IgA generated by vaccination seemed to be generally cross-reactive to spike proteins across the entire *coronaviridae* family, rather than preferentially specific to the vaccine’s cognate antigen. This suggests that prior exposure to endemic human coronaviruses (hCoV) may have generated a memory pool of mucosal B cells. These memory B cells may have been stimulated with our mucosal SARS-CoV-2 vaccine and elicited cross-reactive IgA responses that recognize various human alpha- and betacoronaviruses. Indeed, this has been seen with infection of SARS-CoV-2 but not with the mRNA vaccines (14).

We also showed that the surrogate neutralizing activity persisted past 180 days in the majority of responders, with levels still remaining above pre-vaccination. A recent SARS-CoV-2 study showed mucosal IgA responses correlate to protection from infection; subjects in which lower IgA responses were observed had increased susceptibility to breakthrough infection (15). The authors from the same paper have also concluded that long lasting IgA responses should be the goal of new COVID-19 vaccines (15). It is estimated that IgA can be up to 10 times more neutralizing than IgG and dimeric IgA can be up to 15 times more neutralizing than it’s monomeric counterpart (12) (8), and thus small increases may still be potent. It has been shown that serum from unvaccinated individuals who contracted the omicron variant does not efficiently neutralize other variants. Therefore, generation of mucosal IgA that is cross-reactive could help prevent infection with omicron and other variants (16). The ability to stimulate cross-reactive memory pools of mucosal cells and increase cross-reactive secretory IgA could make VXA-CoV2-1, or future iterations of the product, an attractive pan-coronavirus vaccine candidate.

A limitation of this study was the size of the trial. This Phase I open label clinical trial was run at the start of the pandemic and was useful for demonstrating the immune response profile after a single dose; but was not designed to look at prime-boost in significant numbers or to measure efficacy. Another major limitation is that this vaccine was tested before mRNA vaccines were readily available. Currently, the majority of people in the United States have been immunized with an mRNA vaccine, and it’s unclear how this vaccine would pair with such a population. Given that mRNA vaccines induce potent, but transient serum responses, it is hopeful that this vaccine would complement the mRNA vaccines. A recent study by Mao and colleagues has shown that mucosal boosting can significantly improve cross-reactive immune responses in mice (17).

In conclusion VXA-CoV-2 was well tolerated and induced cross-reactive neutralizing mucosal IgA responses that could persist up to 180 days in the majority of responders. This was the first oral vaccine against SARS-CoV-2 to demonstrate both tolerability and immunity in humans. Subsequent studies will evaluate dose and different antigen formats, as well as efficacy in larger human studies.

## Methods

### Clinical protocol and enrolment

Initially, 5 sentinel subjects were enrolled into Cohort 1 and were immunized with low dose (1×10^10^ IU ±0.5 log) VXA-CoV2-1. Enrollment criteria can be found in the appendix. These subjects recorded solicited adverse events (AEs) daily for 7 days post vaccination. Subjects returned to the site on Day 2 and Day 8 to have safety assessments and samples collected for evaluation of immunogenicity. The study Safety Monitoring Committee (SMC) reviewed the safety data from the sentinel Cohort 1 subjects through their Day 8 visits (7 days post vaccination) which included review of solicited symptoms of reactogenicity, safety lab test results and unsolicited AEs. Upon completion of the SMC review and their recommendation to proceed, enrollment of the remaining low dose, 15 subjects in Cohort 2, and subsequently subjects in Cohort 3 with the high dose (5×10^10^ IU ±0.5 log) of VXA-CoV2-1 was initiated. All subjects in the study followed the same schedule for assessments and sample collection as the sentinel subjects through Day 8. Following Day 8, subjects returned to the site on Day 15 and Day 29 at which they had safety assessments and samples collected for immunogenicity. On Day 29, Cohort 1 sentinel subjects had pre-vaccination safety tests and assessments to determine eligibility to continue with the second vaccination. All subjects entered the long-term safety follow-up period 28 days after their last vaccination, either Day 57 in Cohort 1, or Day 29 in Cohorts 2 and 3. During the safety follow-up period subjects were monitored for safety via monthly calls as well as site visits at Months 6 and 12 where samples to assess for duration of immune responses were also collected.

Nasal swabs, saliva and serum samples were taken from convalescent subjects 1-8 months post confirmed SARS-CoV-2 infection at a single site. All subjects gave informed consent.

### Safety Assessment

Safety was assessed via physical exams, vital signs and safety laboratory tests collected at all predefined site visits. Solicited and unsolicited AEs were collected from time of initial vaccination and assessed by the PI for severity and relatedness to study vaccine. The Safety Monitoring Committee (SMC) oversaw the safety of the study and reviewed safety data from the low dose cohort before initiation of dosing in the high dose group (Cohort 3). Solicited AEs (reactogenicity) were collected with the aid of a 7-day solicited symptoms diary card. Unsolicited AEs (all other clinical AEs) were collected through 28 day post last dose. SAEs, MAAEs and COVID 19 were collected through 1 year post first vaccine dose.

### Endpoints

The primary endpoint for this study was safety. The secondary endpoint, immunogenicity, was assessed through the active phase (Day 29 or Day 57), primarily by measuring IgA and IgG in nasal, serum and saliva. Long-term durability was assessed in subjects that were available. Additional immunological endpoints investigated included T cell responses in PBMCs and immune cell phenotyping (manuscript in preparation).

### Statistics

Statistical analyses were performed using GraphPad Prism v9 software. The method used for determining significance was Mann-Whitney test. *P* values of ≤ 0.05 were considered significant. Error bars are standard error of the mean (SEM).

### Study Approval

The study was conducted in accordance with applicable Good Clinical Practice guidelines, the United States Code of Federal Regulations, and the International Conference on Harmonization guidelines. IRB approval was obtained from Aspire IRB (Seattle/WA; AAHRPP accredited) before study specific screening and enrollment of subjects. Informed consent was obtained from all subjects after discussion of the study procedures and potential risks.

### SARS-CoV-2 Vaccine

VXA-CoV2-1 is a rAd5 vector containing full-length SARS-CoV-2 S gene under control of the CMV promoter and full-length SARS-CoV-2 N gene under control of the human beta-actin promoter. The published DNA sequences of SARS-CoV-2 publicly available (Genbank Accession No. MN908947.3) were used to create recombinant plasmids containing transgenes cloned into the E1 region of Adenovirus Type 5 (rAd5), using the same vector backbone used in prior clinical trials for oral rAd tablets (2). GMP drug substance was produced in HyClone Single Use Bioreactors (GE Healthcare Life Sciences) at Kindred (Burlingame, CA). Purification was performed by ion exchange chromatography, followed by buffer exchange. Purified vector was mixed with excipients, lyophilized, and then tableted at Vaxart using microcrystalline cellulose (FMC) and starch (Colorcon) as tableting bulk. Tablets were enteric coated with Eudragit L100® (Evonik Industries) using a Vector Hi-Coater system (Vector Freund). The final product was released in one lot, and titered by standard IU assay. Placebo was prepared as similarly sized and shaped tablets containing 250 mg of microcrystalline cellulose.

### Measurement of vaccine-induced IgA antibodies in mucosal samples

Nasal samples were collected according to the package insert for the Nasosorption FX-I /SAM devices (Mucosal Diagnostics) and extracted by centrifugation in a 1X PBS with 0.02% azide solution. Saliva samples were collected by centrifugation after having each subject chew on a cotton swab for 45sec and transferred to a salivette (Sarstedt).

IgA was measured with a SARS-CoV-2 specific (V-plex panel 2) Mesoscale Discovery (MSD) immunoassay. Plates were blocked, washed, and incubated with sample and detection antibody according to the manufacturer’s instructions. Samples were diluted at 1:10 and 1:100 in Diluent-100. Plates were read on a Meso QuickPlex instrument. Sample antibody concentrations were reported in arbitrary units (AU) per ml as calculated from a standard curve supplied with the kit. Due to the variability in sampling the human mucosa, samples were normalized using an ELISA for the detection of total IgA. Briefly, purified anti-human IgA mAb MT57 (Mabtech) was coated onto 96-well Maxisorp plates (Thermo Fisher) at 2μg/ml in carbonate-bicarbonate buffer and incubated overnight at 4 °C. Plates were washed with 1X PBS + 0.1% Tween-20 (PBST) and blocked with PBST + 1% BSA for 1hr at room temperature (RT). Following a wash step, saliva and eluted nasal samples were diluted 1:100 in PBST and serially diluted 1:3 down the plate. Human IgA (Sigma-Aldrich) was used to create a standard curve starting at 200ng/ml and serially diluted 2-fold 7 times. The samples and standards incubated for 2hrs at RT. After a wash step, a 1:1000 dilution of anti-human IgA mAb MT20 ALP conjugate (Mabtech) was added to the nasal samples and incubated at RT for 1hr followed by washing. The plates developed for 1hr in the dark with pNPP substrate (Mabtech) and ODs were measured at 405nm with a Spectra Max M2 microplate reader. The concentration of total IgA in human mucosal samples was generated by a standard curve. Samples were normalized per ng of total IgA. Data analysis was performed in Graphpad Prism.

### Surrogate Viral Neutralizing Test

Surrogate neutralizing activity in nasal and saliva samples was measured with a SARS-CoV-2 specific (panel 2) Mesoscale Discovery (MSD) immunoassay. Plates were blocked, washed, and incubated with sample and detection antibody according to the manufacturer’s instructions. Samples were diluted from neat to 1:50 and 1:400 in Diluent-100. Plates were read on a Meso QuickPlex instrument. Percentage lysis was calculated by the reduction in signal over the negative control samples. Sample neutralizing antibody concentrations were reported in arbitrary units (AU) per ml as calculated from a standard curve supplied with the kit.

## Data Availability

All data produced in the present work are contained in the manuscript

## Author Contributions

SJ, MC, SJG, CJM & SNT designed the clinical studies. CIM, CBJ & SG performed the immunological experiments. SJ analyzed the immunological data. NP & EGD provided the vaccine seed material. SJ, SJG, and SNT wrote the manuscript.

## Acknowledgements

Thank you to all the volunteers in the clinical trial and convalescent study.

## Figure Legends

**Supplementary Figure 1:**
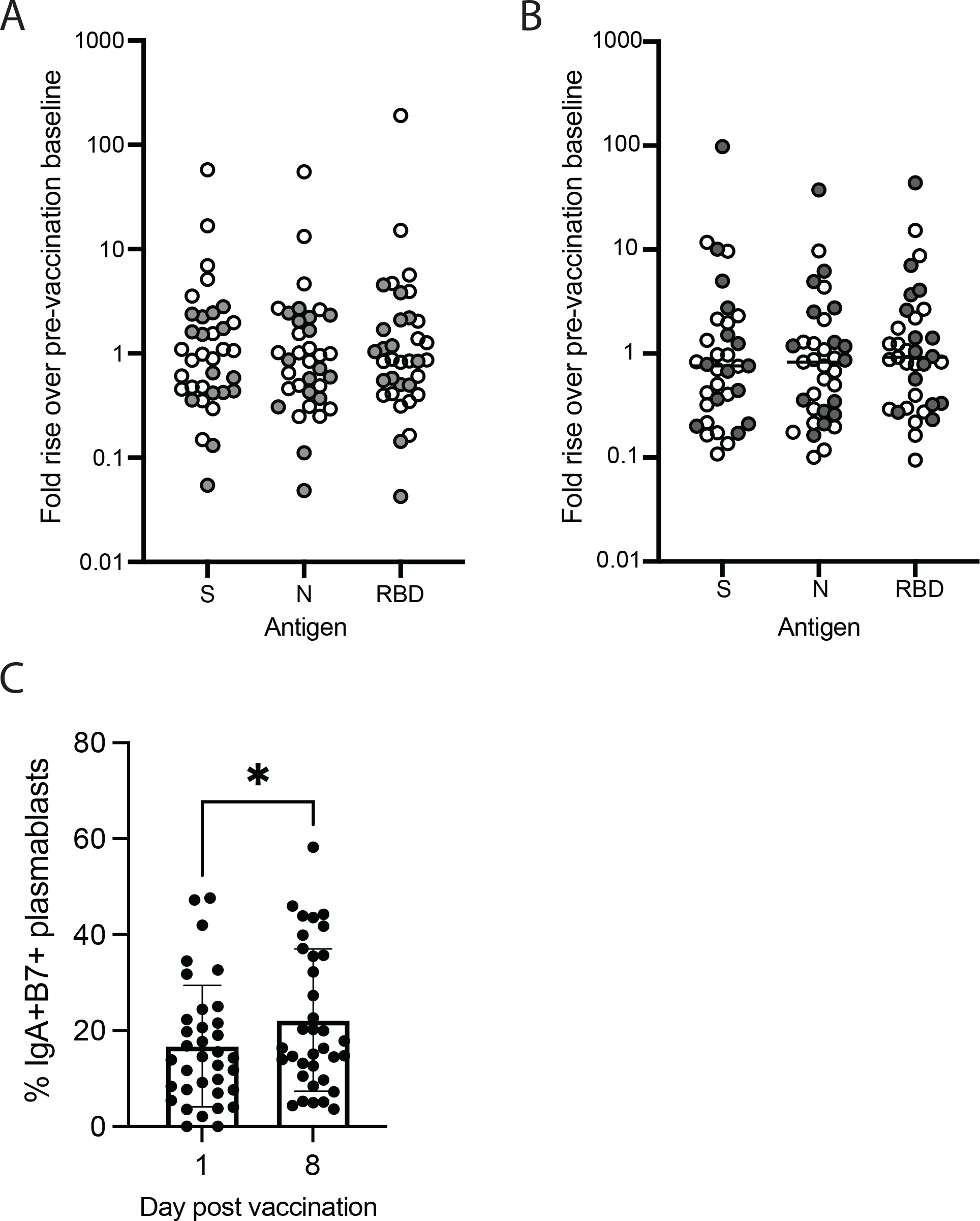
IgA responses stratified by dose. (A-B) IgA was measured using SARS-CoV-2 specific MSD assays in nasal (A) or saliva (B). (C) Frequency of IgA+B7+ plasmablasts as measured via flow cytometry

**Supplementary Figure 2:**
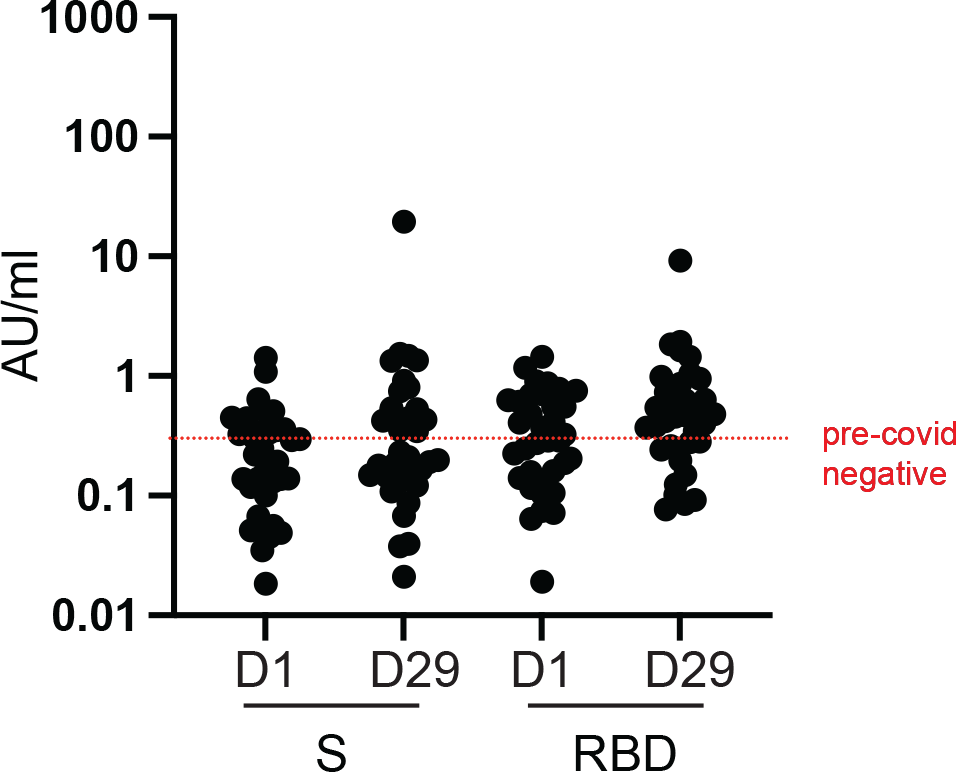
Nasal IgG responses. IgG was measured in the nasal swabs via SARS-CoV-2 MSD. Graph shows AU/ml with. red line representing pre-2019 negatives.

### Appendix

|  |  |
| --- | --- |
| **Eligibility Criteria** | <u>Inclusion Criteria</u><br>1. Male or female between the ages of 18 to 54 years, inclusive.<br>2. Negative for SARS-CoV-2 infection at the time of screening<br>3. In general good health, without significant medical illness, based on medical history, physical examination, vital signs, and clinical laboratories (CBC, chemistry, and urinalysis) as determined by the Investigator.<br>4. Demonstrates comprehension of the protocol procedures and is able to provide written informed consent.<br>5. Available for all planned visits and willing to complete all protocol defined procedures and assessments (including ability and willingness to swallow multiple small enteric-coated tablets per vaccine dose).<br>6. Body mass index between 17 and 30 kg/m<sup>2</sup> at screening.<br>7. Female subjects must have a negative pregnancy test at screening and before each vaccination <u>and</u> fulfill one of the following criteria:<br>a. At least 1 year post-menopausal;<br>b. Surgically sterile;<br>c. Use of oral, implantable, transdermal or injectable contraceptives for 30 days prior to initial vaccination and until 60 days after the last vaccination;<br>i. A reliable form of contraception must be approved by the Investigator (e.g., double barrier method, Depo-Provera, intrauterine device, Norplant, oral contraceptives, contraceptive patches).<br>d. Not be sexually active (abstinent) or be in a relationship with partner who is sterile (must be discussed with site staff and documented).<br><u>Exclusion Criteria:</u><br>1. Known previous exposure to SARS-CoV-2 or receipt of an investigational product for the prevention or treatment of COVID-19, middle east respiratory syndrome (MERS), or severe acute respiratory syndrome (SARS).<br>2. Is in a current occupation with high risk of exposure to SARS-CoV-2 (e.g., health care workers [HCW] or emergency response personnel [“first-responders”] having direct interactions with or providing direct care to patients).<br>3. Individuals with the following underlying medical conditions who are at higher risk of severe illness from COVID-19:<br>a. Cancer, including history of cancer or treatment within past 3 years (excluding basal cell carcinoma or squamous cell carcinoma) |
|  | b. Chronic kidney disease<br>c. COPD (chronic obstructive pulmonary disease)<br>d. Immunocompromised state (weakened immune system) from solid organ transplant, or other medical condition<br>e. Obesity (BMI $\geq 30$)<br>f. Serious heart conditions, such as heart failure, coronary artery disease, or cardiomyopathies<br>g. Sickle cell disease<br>h. Type 2 diabetes mellitus<br>4. Donation or use of blood or blood products within 4 weeks prior to vaccination or planned donation during the study period.<br>5. Diagnosed bleeding disorder or significant bruising or bleeding difficulties that could make blood draws problematic.<br>6. Any condition that resulted in the absence or removal of the spleen.<br>7. Positive HIV, HBsAg or HCV tests at the screening visit.<br>8. Stool sample with occult blood at screening<br>9. Use of antiviral medications, including anti-retrovirals within 7 days before and for 60 days after vaccination.<br>10. Use of antibiotics, proton pump inhibitors, H2 blockers or antacids within 7 days before and for 60 days after vaccination.<br>11. Use of medications known to affect the immune function (e.g., systemic corticosteroids and others) within 14 days before or for 60 days after vaccination.<br>12. Regular use of nonsteroidal anti-inflammatory drugs, sulfonyleureas, and angiotensin II blockers within 7 days before and for 60 days after vaccination.<br>13. Acute disease within 72 hours prior to vaccination defined as the presence of a moderate or severe illness (as determined by the Investigator through medical history and physical exam). (Assessment may be repeated during screening period.)<br>14. History of drug, alcohol or chemical abuse within 1 year of screening.<br>15. Positive urine drug screen for drugs of abuse at screening (except for previous marijuana use); concurrent or ongoing use of marijuana during the active study period through Day 57.<br>16. Consistent/habitual smoking or vaping within 2 months prior to vaccination. Consistent/habitual smoking is defined as smoking 1 or more pack of cigarettes per day, or vaping more than 90 mg nicotine (30 mL bottle of 3 mg e-juice) per day. |
|  | 17. History of hypersensitivity or allergic reaction to any component of the investigational vaccine or placebo, including but not limited to fish gelatin.<br>18. Administration of any investigational vaccine, drug or device within 8 weeks preceding vaccination, or planned use within the duration of the study.<br>19. Any other condition that in the clinical judgment of the investigator would jeopardize the safety or rights of a subject participating in the trial, would render the subject unable to comply with the protocol or would interfere with the evaluation of the study endpoints. |

## Notes

**Conflict of Interest:** Conflict of Interest: All authors are employees of Vaxart, Inc., and have received stock options and compensation as part of their employment.

### Competing Interest Statement

All authors are employees of Vaxart, Inc., and have received stock options and compensation as part of their employment.

### Clinical Trial

NCT04563702

### Funding Statement

This study did not receive any external funding

